# Bone Densitometry Dataset for Computer Aided Osteoporosis Disease Detection

**DOI:** 10.1101/2025.01.25.24319689

**Authors:** Negin Masnabadi, Abolghasem Sadeghi-Niaraki, Mohammad Karimi, Tamer AbuHmed, Nasrin Azarbani, Soo-Mi Choi

**Affiliations:** Geoinformation Tech. Center of Excellence, Faculty of Geodesy& Geomatics Engineering, K.N.Toosi University of Technology, Tehran, Iran; Dept. of Computer Science & Engineering and Convergence Engineering for Intelligent Drone, XR Research Center, Sejong University, Seoul, Republic of Korea; College of Computing and Informatics, Sungkyunkwan University, Suwon16419, Republic of Korea; Department of Internal Medicine, Assistant professor of Rheumatology, Arak University of Medical Sciences, Arak, Iran

**Keywords:** osteoporosis, clinical data, T-score, BMD, DXA, X-ray

## Abstract

Recently, automatic disease diagnosis based on medical images has become an integral part of digital pathology packages. To create, develop, evaluate, and compare these systems, we need diverse data sets. One of the key features in the diagnosis of bone diseases is measuring bone mineral density (BMD). Most research in this field uses manual methods to directly extract bone image features despite the underlying correlation between diseased and healthy bones, which explains the limited results. Detection of significant changes in bone mineral density (BMD) relies on minimally invasive dual energy x-ray absorptiometry (DXA) scanners. This article presents a collection of bone density test results along with a patient profile called Arak Bone Densitometry Center data. The patient profile includes height and weight and information about the patient, along with photos of the imaging areas. The number of these patients is 3,643, with about 4,020 photos stored next to them. Which can be used to develop automatic disease diagnosis methods and software.

**Dataset:** https://drive.google.com/drive/folders/1HmLTG4GFgB2s4D0×7TTRx8vV_VWY3sW3?usp=sharing

## 1. Summary

The adult human body needs 206 unique structures to withstand the physiological loads that are imposed on it as a result of performing daily activities, each of which has properties that are harmoniously adapted to support the loads imposed on its tissue level. Each bone consists of a spongy trabecular bone network surrounded by a dense cortical shell (Figure.1). The strength of the bone depends on the thickness of the cortical and the porosity significantly. The proportion of each part of the bone is different depending on the placement. Bone size[1], bone shape[2, 3], architecture[4, 5], composition (minerals, matrix, etc.)[6-8] are among the factors that the combined sum of these characteristics determine bone strength.

**Figure. 1.**
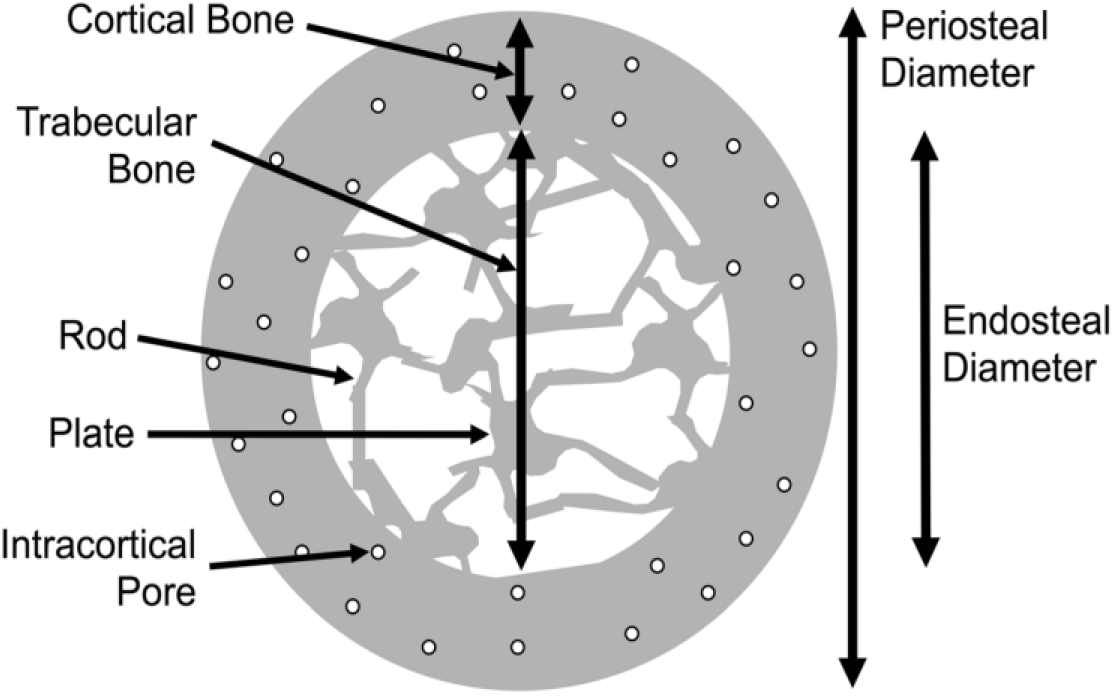
Bone structure

**Figure. 2.**
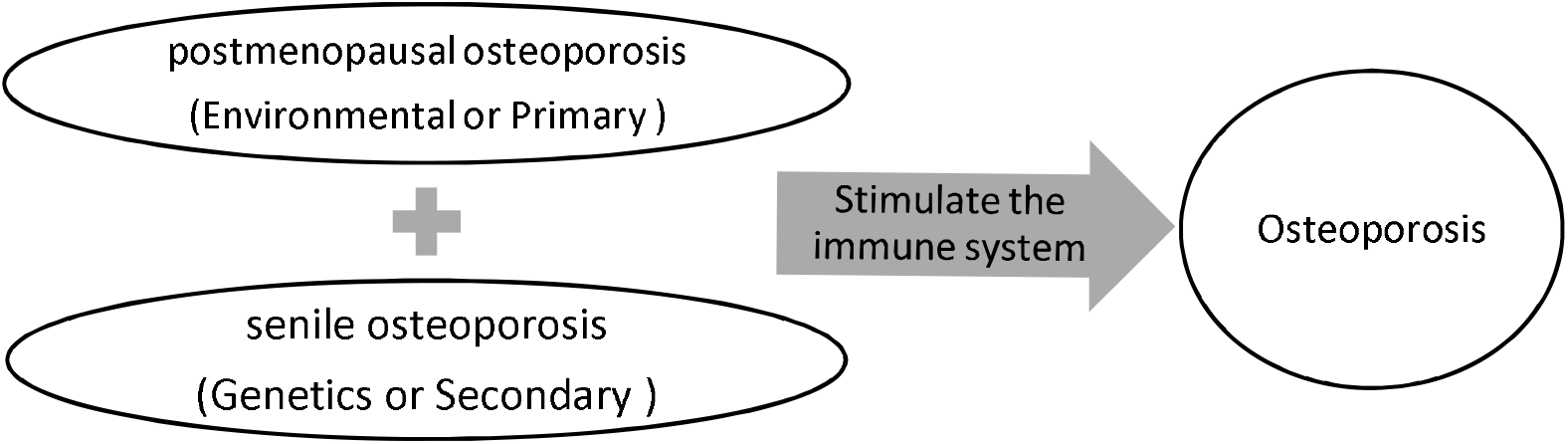
two types of osteoporosis type) causes irritation of the nervous system and ultimately osteoporosis. (Figure.2)

Osteoporosis is a serious public health problem and one of the most common bone diseases in humans[9]. Among the symptoms of this disease, we can mention a decrease in bone density, microarchitectural changes in bone tissue, increased tendency to fracture, and bone fragility, all of which involve the skeleton [9]. The phenomenon of osteoporosis predominantly manifests in women after menopause[10], a subject inundated with various directives and protocols concerning its assessment and control within this demographic. Concurrently, the emergence of male osteoporosis has emerged as an escalating public health quandary, warranting heightened scrutiny and discourse. Osteoporosis was described as: “a systemic skeletal disease characterized by low bone mass and microarchitectural destruction of bone tissue with increased bone fragility and susceptibility to fracture,” a conceptual definition that was adopted several years later by the NIH Community Development Panel was supported Osteoporosis [11].

Considering its cause, osteoporosis can be divided into two categories, the primary category includes people who develop this disease after menopause, and the second category includes people who develop this disease as they age [12] [13]. The cause of osteoporosis of the first type is the cessation of periods in women [14]. But the second type of this disease can be caused by many factors, which are more difficult to diagnose than the first type, and usually the disease of these patients is not diagnosed and includes men and women. Among the reasons that cause the loss of bone density in these people(second type), we can mention the adverse effects of drug therapy, endocrine illnesses, eating disorders, kidney disease, and cancer. In general, it is caused by various illnesses and drug side effects[15]. The cause of each of the two types of environmental (primary type) and genetic (second

Osteoporosis is a significant health challenge, primarily due to the high rates of fragility fractures, which subsequently increase both mortality and morbidity. There is a crucial international debate regarding the adoption of protocols for the diagnosis and management of osteoporosis[16].Currently, the average age of the world population is increasing (old age)[17], this disease is an important public health challenge. Statistics show that 8.9 million fractures are due to osteoporosis, of which more than 1.5 million occurred in the United States [18]. In the United States, it doubled (to more than 3 million). [19]. The highest prevalence of osteoporosis is reported in Africa at 39.5% (95% confidence interval, CI 22.3-59.7) [20].According to published statistics, the incidence of fractures has increased significantly, and this increase can be caused by several factors, including the lack of awareness of doctors, patients, and other health professionals, treatment gaps, and mismanagement in this field. Prevention of the occurrence and early detection of the disease are considered necessary goals[21].

Various methods are available for detecting bone density, each with its own set of advantages and disadvantages. These methods include Dual-energy X-ray absorptiometry (DXA), quantitative computed tomography (QCT), peripheral quantitative CT (pQCT), high-resolution peripheral quantitative CT (HR-pQCT), and ultrasonography (QUS). Among these, DXA remains the most common and reliable technique for assessing bone density. By utilizing the results of these tests, osteoporosis can be categorized into three types: normal, osteopenia, and osteoporosis (Suhas, Satyavaishnavi et al., 2012). DXA, which stands for Dual-energy X-ray absorptiometry, is widely used due to its accuracy and precision in measuring bone mineral density (BMD). It works by passing low-dose X-ray beams through bones, measuring the amount of radiation absorbed by bone tissue. This method is particularly effective in diagnosing osteoporosis and assessing fracture risk. Quantitative computed tomography (QCT) is another technique that measures bone density by analyzing CT scans. Unlike DXA, QCT provides three-dimensional images of bone density, offering detailed information about bone structure and density distribution. However, QCT is less commonly used than DXA due to its higher cost and radiation exposure. Peripheral quantitative CT (pQCT) and high-resolution peripheral quantitative CT (HR-pQCT) are specialized forms of CT imaging that focus on peripheral bones, such as those in the forearm and lower leg. These techniques are valuable for assessing bone density in specific regions of the body and are often used in research settings to study bone health and related conditions. Ultrasonography (QUS) is a non-invasive technique that uses sound waves to measure bone density at peripheral sites, such as the heel or finger. While QUS is portable and radiation-free, its accuracy may vary depending on the operator’s skill and the quality of the equipment. Despite these limitations, QUS can be a useful screening tool for identifying individuals at risk of osteoporosis, especially in community healthcare settings. Using the results of these tests, the types of osteoporosis are divided into three categories (1) normal (2) osteopenia (3) osteoporosis [22]. In Figure 3, three categories of bone density can be seen.

**Figure. 3.**
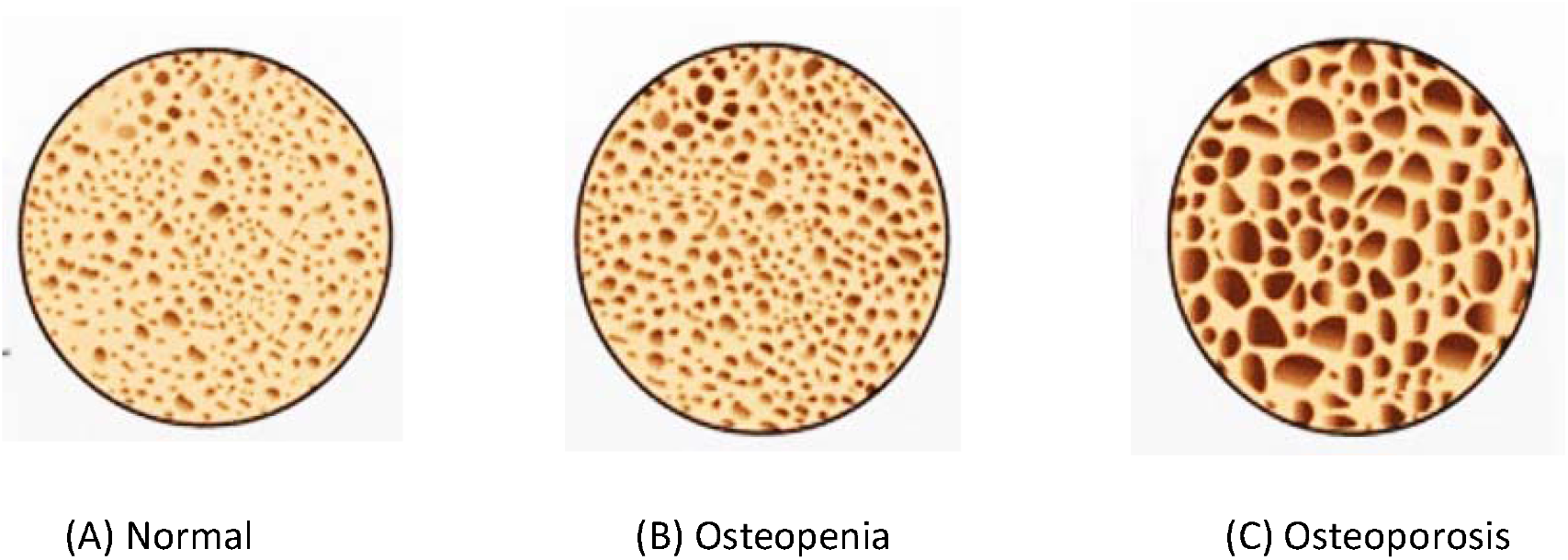
Bone density [23]

In our extensive review of literature pertaining to osteoporosis, it became increasingly evident that a comprehensive and contemporary dataset is conspicuously absent within the realm of osteoporosis research. Table 1, which offers a comparative analysis of several scholarly articles, underscores this glaring gap in the field. It reveals that a substantial portion of the available data remains unpublished and inaccessible to the broader scientific community, thereby hindering progress in understanding and addressing osteoporosis effectively. Of particular significance is the observation that existing datasets lack integration of patient photographs, a notable deficiency in current research efforts. This absence not only limits the depth and breadth of available data but also overlooks potential insights that visual documentation of patients’ conditions could provide. Addressing this shortfall could potentially enhance diagnostic accuracy, treatment efficacy, and overall research outcomes in osteoporosis studies. Moving forward, there is a pressing need to develop and disseminate comprehensive datasets that encompass a broader range of patient information, including visual documentation. By adopting a more inclusive approach to data collection and sharing, researchers can foster collaboration, accelerate discoveries, and ultimately improve outcomes for individuals affected by osteoporosis.

**Table 1.** Comparison table of articles dataset.

| Reference | year | Country | number of cases | number of images | patient information | dataset type |
| --- | --- | --- | --- | --- | --- | --- |
| [24] | 2014-2017 | China | 808 | 1,616 | Yes | private |
| [25] | 2016-2017 | Iran | 1,990 | None | No | Private |
| [26] | 2000-2003 | Iran | 4,229 | None | Yes | Private |
| [27] | 2013-2020 | Korea | 547 | 2,696 | No | Private |
| [28] | 2012-2016 | America | 9,330 | None | Yes | Public |
| [29] | 2017-2018 | China | 2,711 | None | Yes | Private |
| [9] | 2018 | Unknown | not access | not access | No | Private |
| [30] | 2020-2021 | Maqedonise | 657 | None | No | Private |
| Our dataset | 2019-2023 | Iran | 3643 | 4020 | Yes | Public |

The significance of this research lies in its contribution to the provision of a comprehensive dataset for future investigations in the field of osteoporosis. The dataset was meticulously compiled from the Arak bone density measuring center utilizing the DXA method, encompassing a total of 3,644 patients. Spanning from May 9, 2019, to January 15, 2023, the dataset encapsulates a wide array of demographic and clinical information, as outlined in Table 2. This rich repository of data holds immense potential to fuel advancements in osteoporosis research, facilitating deeper insights into the disease’s prevalence, progression, and treatment outcomes.

**Table 2.** The database specifications.

|  |  |
| --- | --- |
| Subject | bone density |
| Modality | Bone density measure(DXA), bone image |
| Type of data | CVC, PNG |
| Data source location | Dr.Tasorian - Dr.Azarbani bone densitometry clinic – Arak |
| Data accessibility | https://drive.google.com/drive/folders/1HmLTG4GFgB2s4D0x7TTRx8vV_VWY3sW3?usp=sharing |
| Experimental factor | All patients were placed in one of the osteoporosis classes |

Continuing with our research endeavor, we will proceed to unveil the entirety of the dataset. Part 2 of our study, titled “Osteoporosis Bone Density Measurement Dataset,” will comprehensively delineate the dataset, elucidating the information contained within each segment, including tabular data and accompanying images, which will be annotated for clarity and relevance. Subsequently, the following section will delve into the methodologies employed for data collection and dataset generation, shedding light on the meticulous processes undertaken to ensure data accuracy and integrity. In the subsequent phases of our research, specifically the 4th and 5th parts, we will embark on a rigorous analysis of the dataset, extracting valuable insights and drawing meaningful conclusions. Furthermore, we will outline avenues for future research endeavors, identifying areas warranting further exploration and potential avenues for enhancing our understanding of osteoporosis and its management strategies.

## 2. Osteoporosis Disease Bone Densitometry Dataset

### 2.1 Ethics statement

In pursuit of ethical rigor and adherence to legal mandates, the authors of this article meticulously observed Iran’s patient rights law throughout the course of their research endeavors. The collected data is fully anonymized, and no personal identifiers such as names, addresses, or contact details were included. This comprehensive approach ensured that every step undertaken, from data collection to analysis, respected the fundamental rights and dignity of the patients involved. Central to this ethical framework was the rigorous scrutiny and approval process facilitated by the ethics committee of the Arak Bone Density Measurement Center, an esteemed body tasked with safeguarding the welfare of research participants. The formal approval granted by the ethics committee not only signifies compliance with regulatory protocols but also underscores a commitment to upholding the highest ethical standards within the medical community. This endorsement provides assurance to both the academic community and the broader public regarding the integrity and ethical soundness of the dataset under examination. Furthermore, the approval for publication serves as a testament to the dataset’s scientific validity and ethical robustness, affirming the authors’ dedication to transparency and responsible data dissemination.

### 2.2 data description

The osteoporosis data set under consideration was meticulously gathered over a span of nearly four years, from May 9, 2019, to January 15, 2023, from Dr.Tasorian-Dr.Azarbani’s Bone Densitometry Center in Arak, Iran. Utilizing cutting-edge technology, specifically the DXA model (S/N 301654), renowned for its precision in American heological density assessment, the center adeptly imaged and measured the bone density of 3,643 patients. Notably, this device transcends mere density measurement, boasting capabilities for fracture risk assessment and other diagnostic evaluations through the innovative utilization of dual-energy X-rays. The patient demographic within the data set skews predominantly towards females, comprising 3,232 individuals, while 411 males are also represented(Figure.4). The distribution of bone density conditions among the patients reveals compelling insights: at the hipneck region, 28% exhibit normal density, 52% fall into the osteopenia category, and 20% are diagnosed with osteoporosis. Similarly, within the spine area, the distribution unfolds as 26% normal, 44% osteopenia, and 30% osteoporosis. Interestingly, the hip area delineates a distribution of 50% normal, 41% osteopenia, and 9% osteoporosis. Intriguingly, each patient’s record within the data set is meticulously preserved, ensuring at least one image per individual for comprehensive analysis and future research endeavors.

**Figure. 4.**
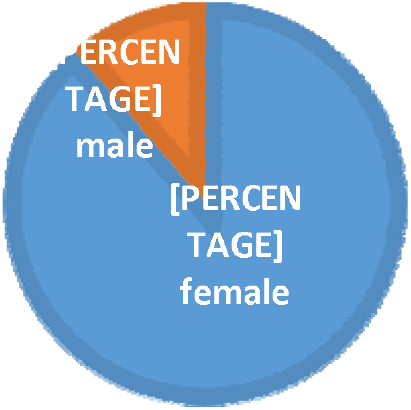
Data gender chart

**Figure. 5.**
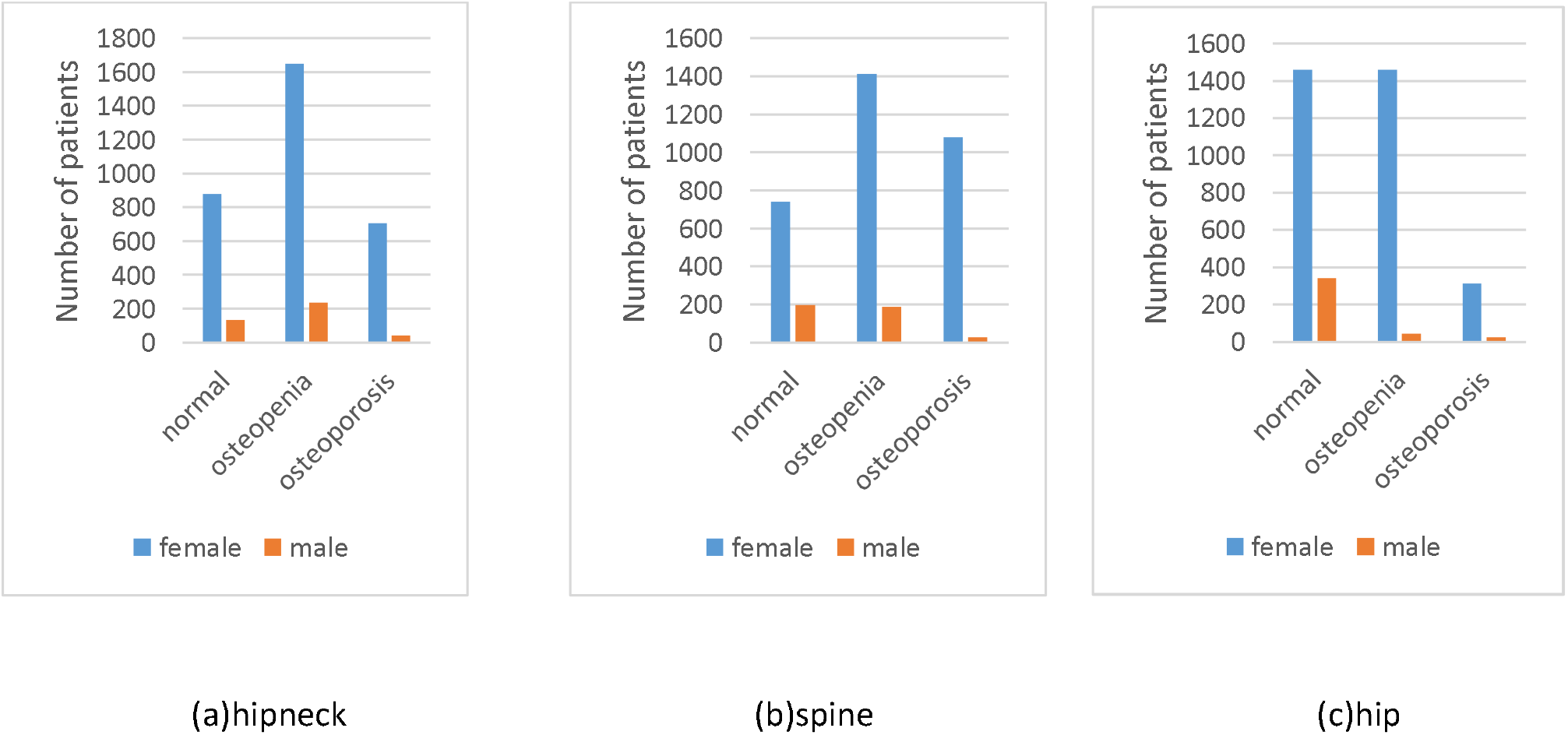
Grouping of data in osteoporosis presented data set

In delineating the dataset’s structure, it becomes evident that it comprises two distinct components, each serving a unique analytical purpose(Figure. 6). The initial segment entails tabular data, meticulously organized to encapsulate essential patient information, facilitating comprehensive statistical analysis and trend identification. This structured data format enables researchers to discern patterns and correlations within the dataset, thereby unraveling valuable insights into the prevalence and characteristics of osteoporosis within the studied population. Conversely, the second component encompasses a repository of images extracted from the software database, offering visual representations of bone density assessments. These images serve as crucial diagnostic aids, affording clinicians and researchers the opportunity to visually inspect and interpret bone density measurements with precision and accuracy. Table 3 shows an example of each group (normal, osteopenia and osteoporosis) from the data set. By coupling quantitative data analysis with visual representations, researchers can attain a holistic understanding of osteoporosis prevalence and severity, fostering advancements in diagnosis, treatment, and preventative strategies.

**Figure. 6.**
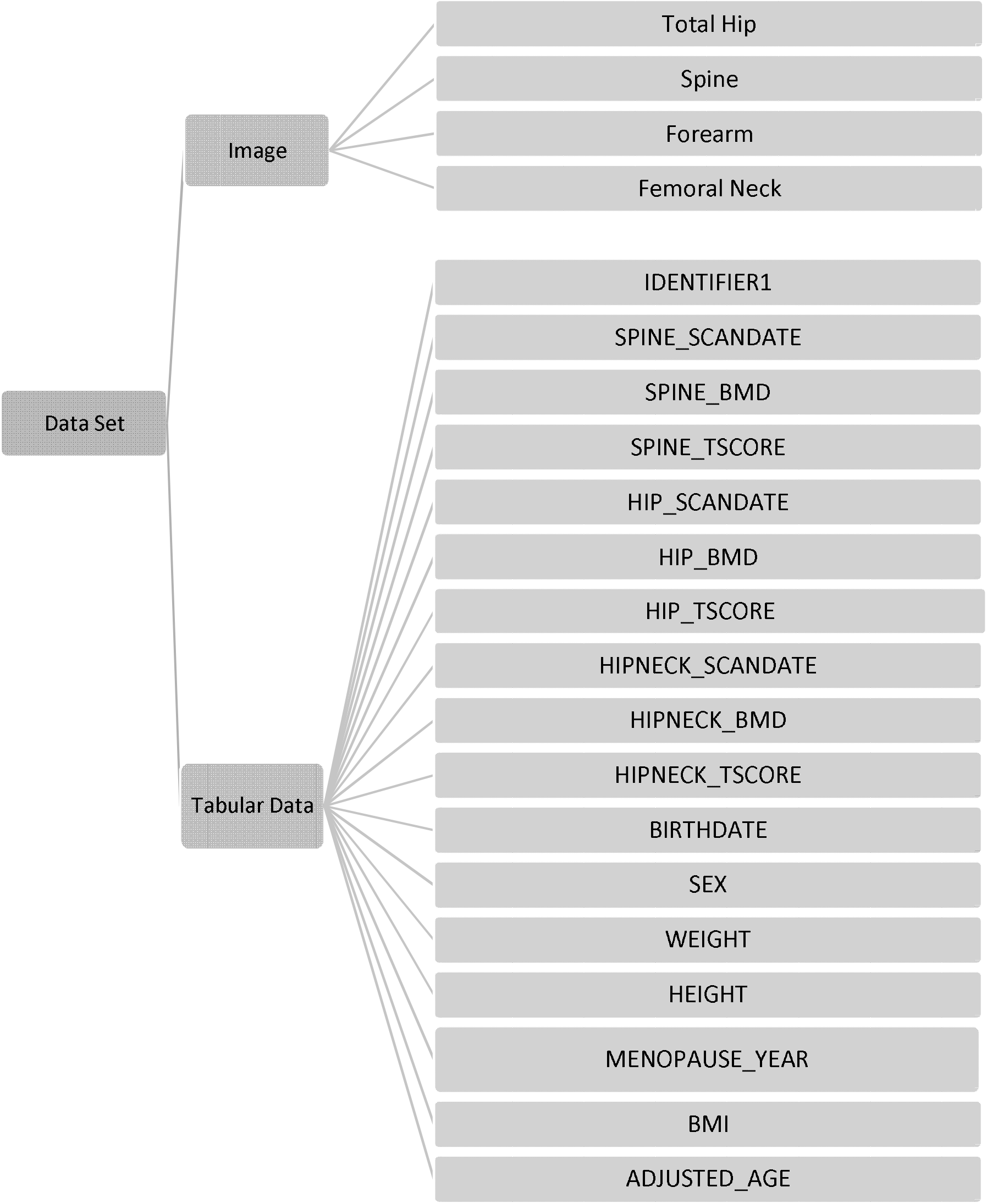
categories of collection

**Table 3.**
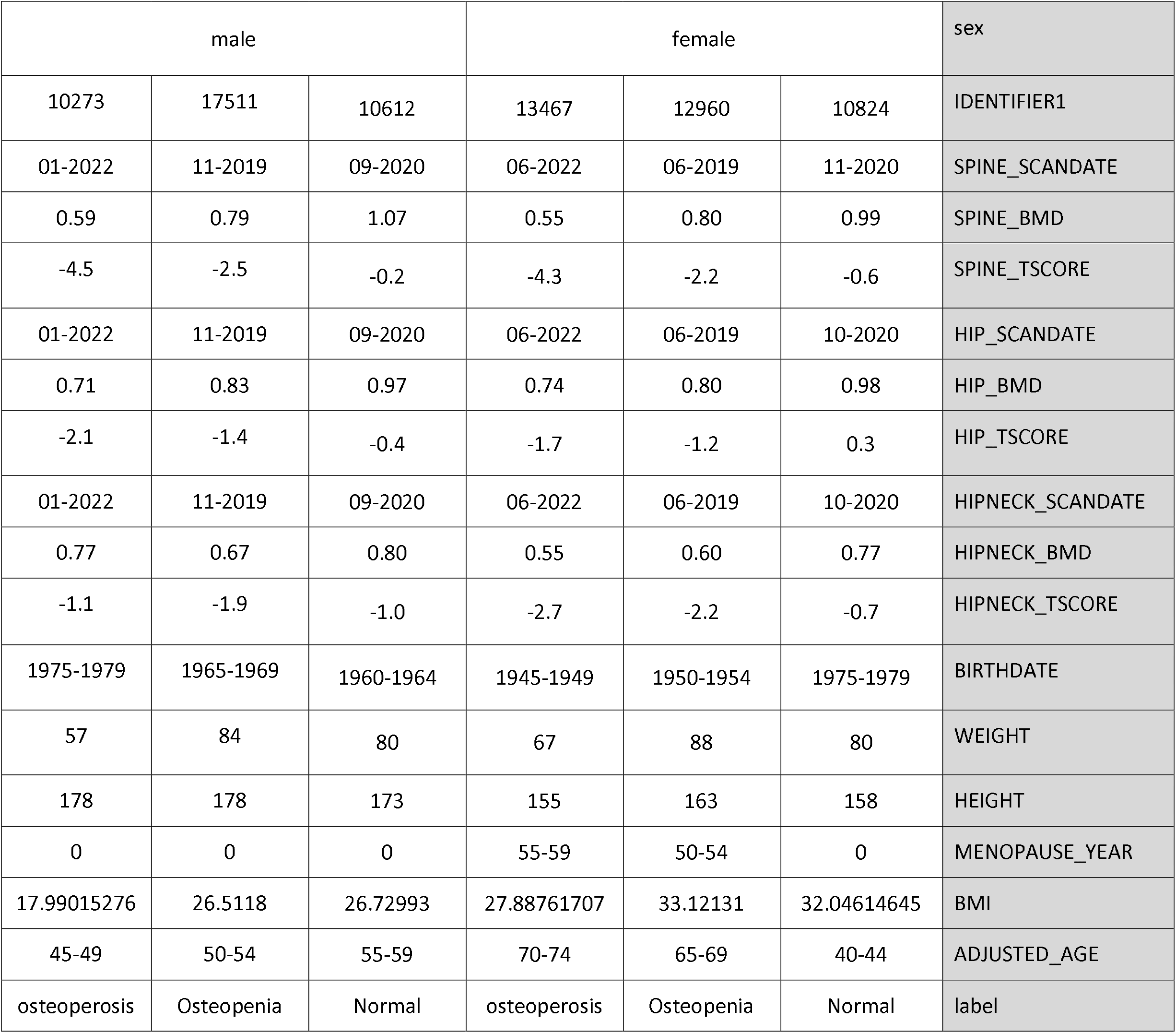
Example of a dataset.

#### 2.2.1. Table information

The information table extracted from the answers of the tests includes lines where each line represents the test answer of one person. This table has 17 columns, and we will discuss the meaning of each row below (table-4).In the table below, we will explain the meaning of that column, standard deviation, and its maximum and minimum value or its number. As it can be seen in the table, the date of taking photos from each area has been recorded, and most of the photography was done at the same time, but in some cases, taking photos from one area is postponed to a later time.

**Table 4.** The information table.

| Table 4 – The information table |  |  |  |  |
| --- | --- | --- | --- | --- |
| Row | Name of column | Definition | Mean ± SD | MIN-MAX or COUNT |
| 1 | IDENTIFIER1 | An anonymized patient identifier (primary key) used exclusively within the research group to link each patient to their photos. | - | - |
| 2 | SPINE_SCANDATE | Date of taking test from patients | - | 10-2019 - 01-2023 |
| 3 | SPINE_BMD | This BMD is for the spine area. | 0.837±0.157 | - |
| 4 | SPINE_TSCORE | This score is obtained by comparing the bone density of the patient's spine with healthy and young people of the same sex. | -1.935±1.435 | - |
| 5 | HIP_SCANDATE | Date of taking test from patients | - | 10-2019 - 01-2023 |
| 6 | HIP_BMD | This BMD is for the hip area. | 0.814±0.143 | - |
| 7 | HIP_TSCORE | This score is obtained by comparing the bone density of the patient's hip with healthy and young people of the same sex. | -1.098±1.134 | - |
| 8 | HIPNECK_SCANDATE | Date of taking test from patients | - | 10-2019 - 01-2023 |
| 9 | HIPNECK_BMD | This BMD is for the neck area. Femoral neck BMD is a strong predictor of hip fracture susceptibility in elderly men and women because it detects cortical bone instability. | 0.676±0.135 | - |
| 10 | HIPNECK_TSCORE | This score is obtained by comparing the bone density of the patient's hip neck with healthy and young people of the same sex. | -1.593±1.1738 | - |
| 11 | BIRTHDATE | Birth year categorized into 5-year ranges | - | 1925-1930 to 2010-2015 |
| 12 | SEX | Gender of patients | - | Female =3,223 , male =411 |
| 13 | WEIGHT | Weight of patients | 69.587±13.344 | - |
| 14 | HEIGHT | Height of patients | 159.091±13.830 | - |
| 15 | MENOPAUSE_YEAR | Menopause is when a person has gone 12 consecutive months without menstruation. This column shows the year of menopause for female patients, categorized into 5-year ranges. | ±6 | 35-40 to 55-60 |
| 16 | BMI | Body mass index (BMI) is a criterion for determining the appropriateness of weight and height. BMI is a person's weight in kilograms divided by the square of height in meters. A high BMI can indicate high obesity. | 28.360±22.704 | - |
| 17 | ADJUSTED_AGE | Age-adjusted test, categorized into 5-year ranges. | 38±203 | 20-25 to 80-85 |

#### 2.2.2. Images

The second part of the data includes photos recorded by the device to measure bone density. In addition to the fact that bone density is measured from the photos, the health of the bones can also be seen. For photography, the patient must lie down in the positions you see in the picture below (Figure.7) so that photography can be done in that area. Photography is usually done from the four AP regions of the spine (L1-L4), femoral neck, entire hip and forearm (Figure.8), which are relatively vulnerable and busy bones. If osteoporosis is detected in any of these areas, the result of the test will be determined.

**Figure. 7.**
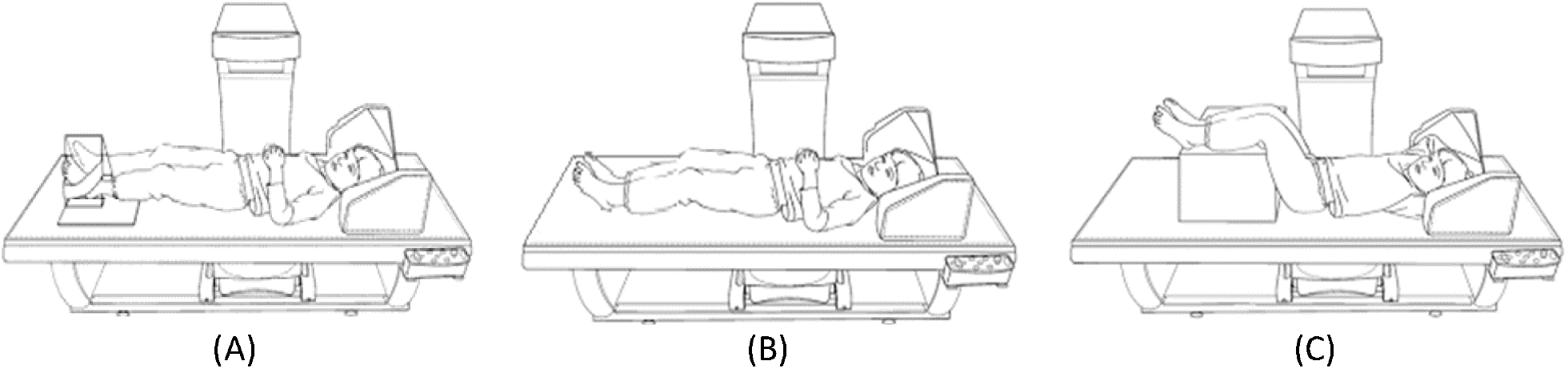
Different positions of the patient to photograph the desired areas

**Figure. 8.**
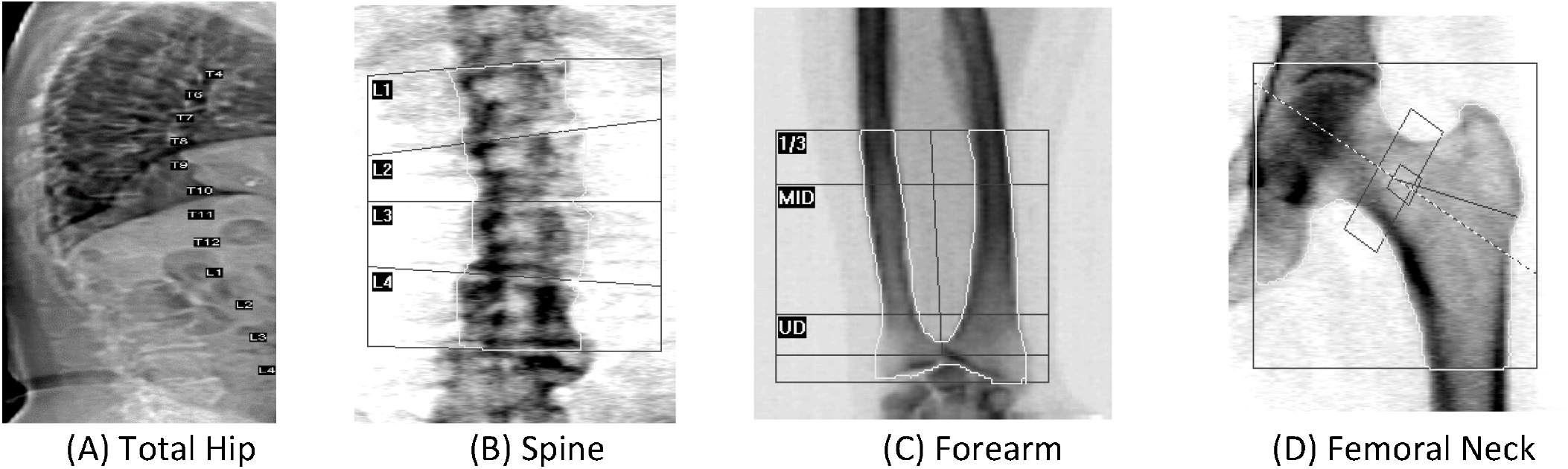
Photographs were taken from different areas

The extracted photos of these patients are 4,020 photos and are stored in a folder called Image. The format of the photos is PNG, and their naming is based on the IDENTIFY1 column, because a code has been selected as the main key for each patient, for patients whose number of photos was more than one, for the next photos, after IDENTIFY1 numbering has been done (Figure.9).

**Figure. 9.**
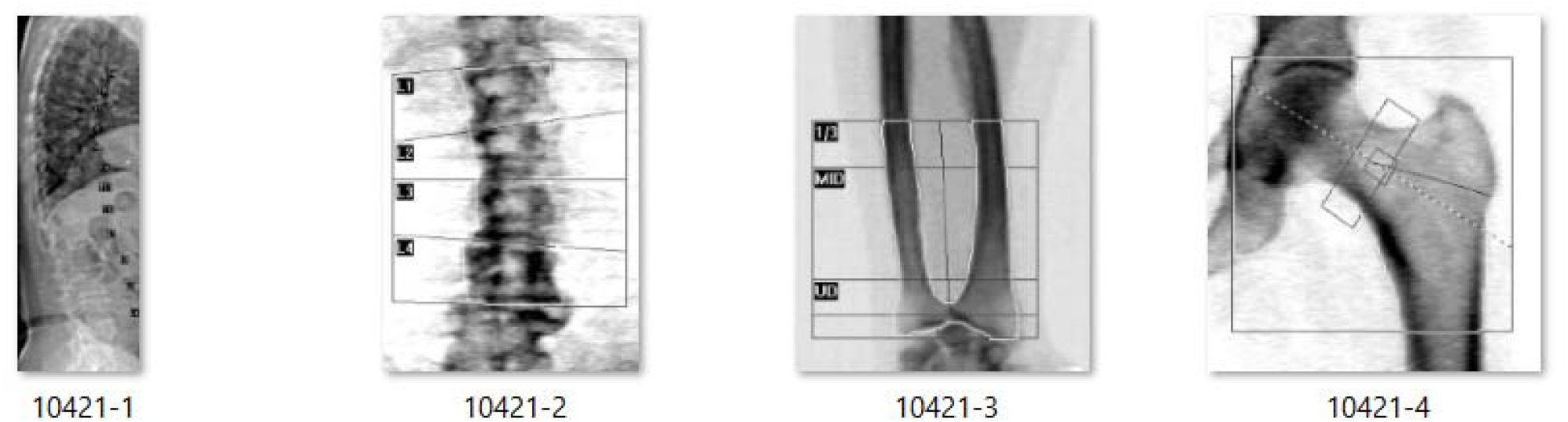
An example of how to name photos

### 2.3 Patient data anotation

First, a photograph is taken of each part of the patient’s body, which is determined by the doctor. Then the software of the device automatically determines the limits that are suitable for detecting the density of that area, which must be confirmed by the operator. The operator can correct the areas if they are wrong. In the photographs taken, these areas are marked as shown in Figures 6 and 7 and then labeled. These areas in the bones are places that have the highest pressure, so they must be stronger (Figure.10). In the spine, the last four vertebrae, in the forearm, one third of the bone and in the middle of the bone and unidirectional, in the Femoral it is measured in the middle of the depression.

**Figure. 10.**
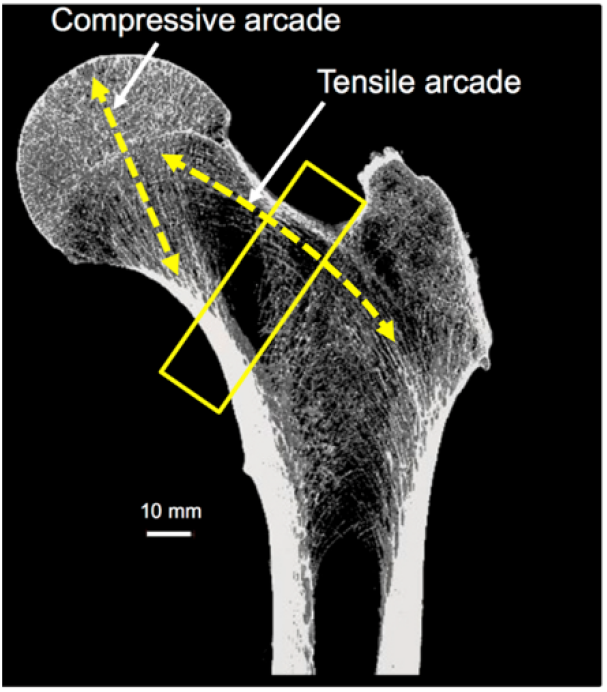
An example of how to name[31]

## 3. osteoporosis data capture method

Osteoporosis is one of the diseases that usually have no symptoms [32]. Doctors recommend all women over 65 and men over 70 and other groups with the following characteristics:

- Postmenopausal women
- Thin people (body mass index less than 18.5)
- Corticosteroids users
- Users of anti-accelerating or seizure medications
- Patients with spinal cord injury or any disease that leads to immobility
- Those who have gone through menopause before the age of 45

Refer to a specialist doctor. In people whose doctor suspects this disease, osteoporosis should be confirmed through bone density measurement. There are different methods for measuring bone density, which we will briefly discuss below.

### DXA

The double X-ray absorption method is one of the most accurate methods used to measure bone density. The World Health Organization has introduced this method as the best densitometry method and presented its classification based on this method. This method divides people by defining T-score [33]. In Table 5, you can see this classification using the T-score. In this method, the three important components of bone, fat tissue, and non-fat tissue of the body are evaluated relatively accurately with the amount of radiation absorption. In Figure 11, you can see example of the normal group, in Figure 12, example of osteopenia and in Figure 13, you can see example of osteoporosis.

**Table 5.** Classification of people using T-score.

| Categories | T score |
| --- | --- |
| (1) normal | At or above -1 |
| (2) osteopenia | Between -1 and -2.5 |
| (3) osteoporosis | At or below -2.5 |

**Figure. 11.**
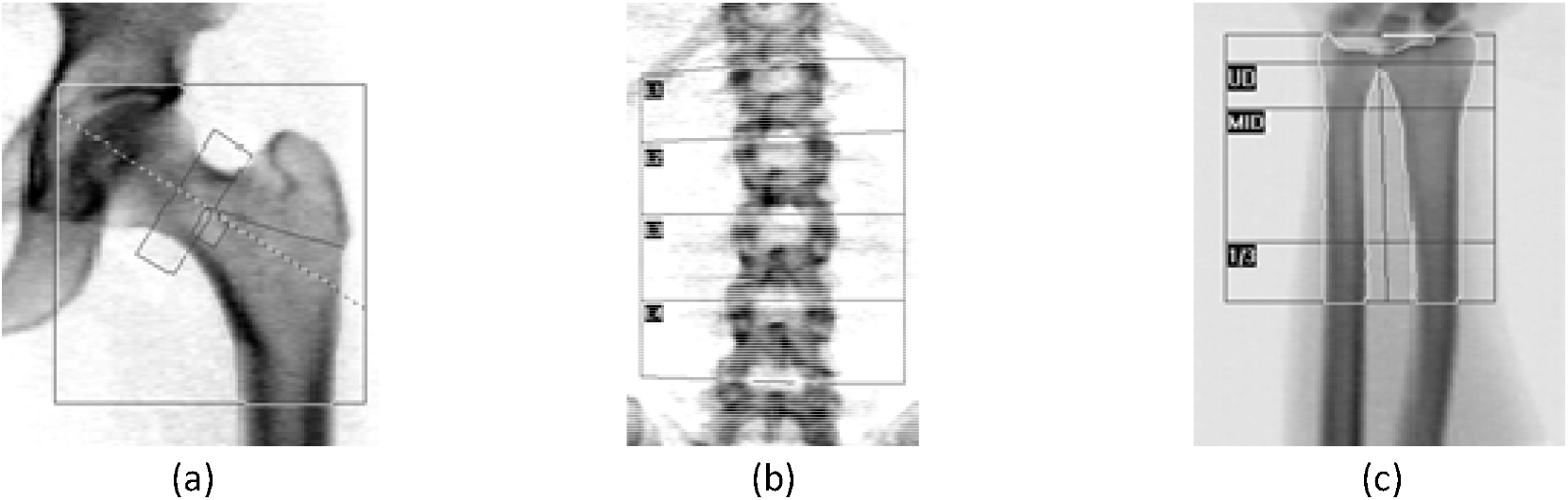
An example of normal data (male):(a) Forearm Neck, (b) Spine and (c) Forearm

**Figure. 12.**
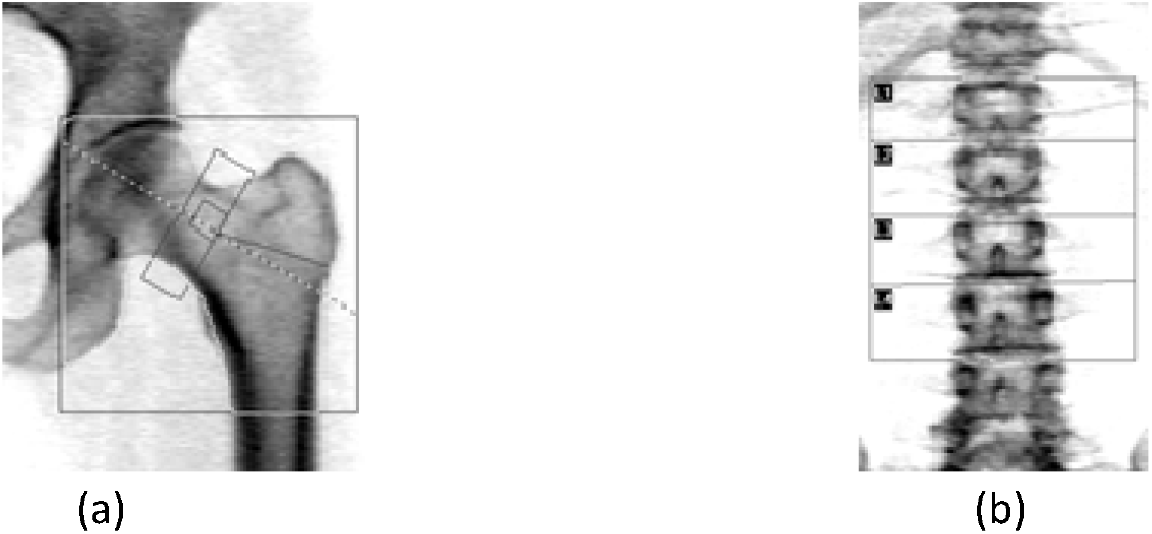
An example of Osteopenia data (Female):(a) Forearm Neck, (b) Spine

**Figure. 13.**
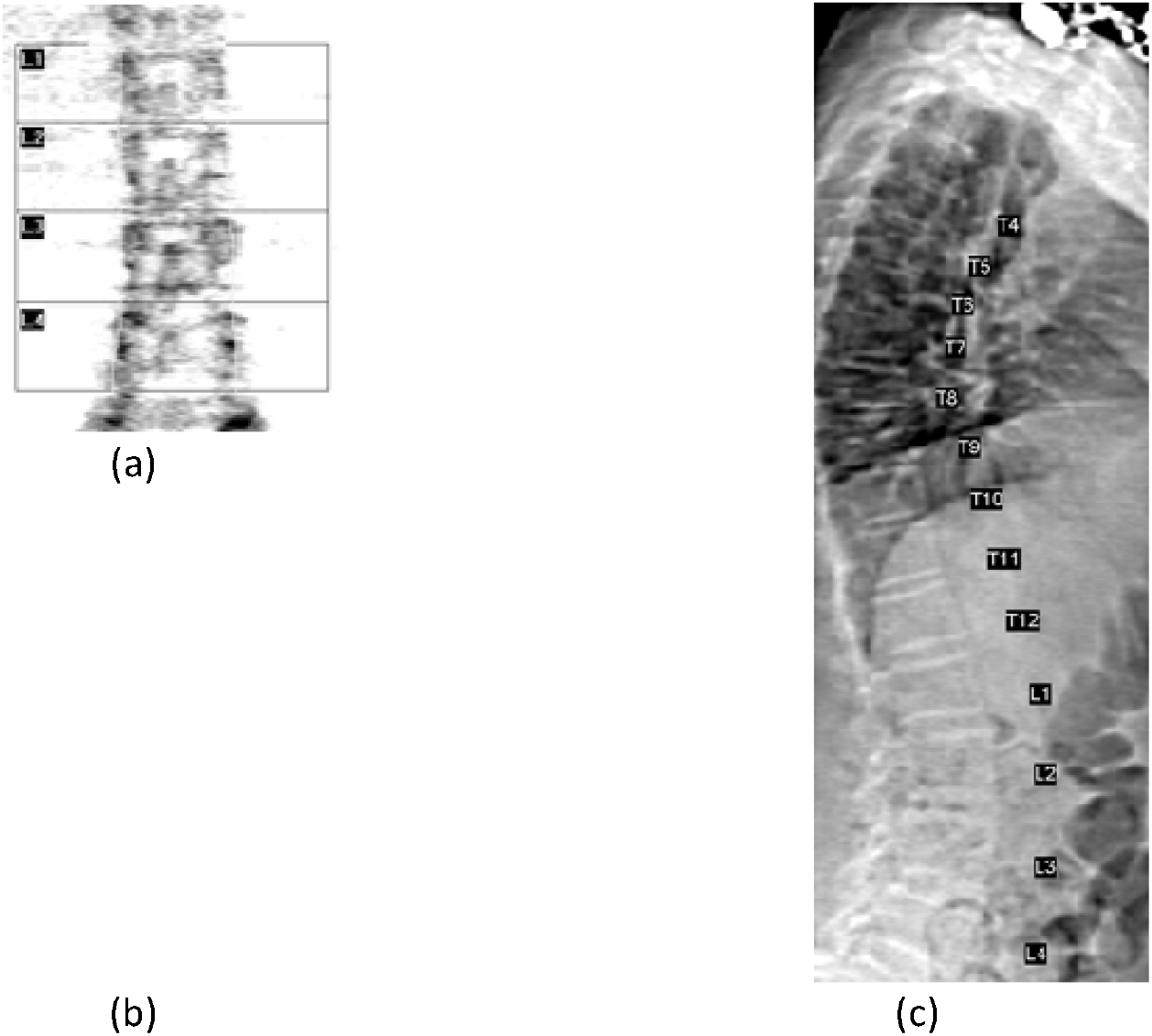
An example of Osteoporosis data (female):(a) Spine, (b) Femoral Neck and (c) Total Hip

### QCT

X-ray quantitative computed tomography or QCT can measure BMD. This device is calibrated using calcium hydroxyapatite, a solid phantom [34, 35]. This value is measured by converting Hounsfield units from CT images to BMD. The radiation dose of this method is higher than DXA, which is one of its disadvantages [36].

### HR-QCT

High-resolution peripheral computed tomography that detects fine bone structures [19]. Using this method, the course of the disease can also be evaluated ;that is, the effects of treatment, bone changes, or the progress of the disease can be observed [37].

### QUC

Quantitative ultrasonometry is an inexpensive method to measure bone density compared to previous methods. This non-invasive method measures the amount of BMD by determining the speed of sound and does not involve any radiation for the patient [38].

## 4. Discussion

This research is one of the first studies that collects and publishes the results of osteoporosis assessment along with the corresponding annotated images.The data collection presented in this article is for the purpose of designing and developing automatic disease diagnosis software and improving the level of knowledge to prevent and improve the condition of patients in this field. Deep learning methods, which are one of the promising methods of classifying medical images, require quality and balanced data sets. Deep learning methods that examine voluminous data sets can help researchers to accelerate the examination of imaging data, including classification and retrieval, and thus achieve better clinical results[39].

Among the problems of the articles reviewed by the authors, we can mention the small number of data or not having enough individual information of sick people. These problems lead to challenging the correctness of the results or evaluating different methods. The volume, quality, and information of the provided data set can meet these needs to some extent. But unfortunately, this dataset also faces challenges, such as the lack of a complete gallery of photos taken from the patient, all of which result in test answers, and the lack of information about the patient’s personal life, such as the location of the place of residence, type of job, the number of children, the history of personal or family illness that accompanied this information leads to better results. Another problem with the presented dataset is the unbalanced number of men and women suffering from this disease. This problem is due to the fact that osteoporosis is more common in postmenopausal women[40], but osteoporosis in men is also considered as a growing public health concern[41]. which can be compensated with techniques to balance and increase. Of course, the fact that one of the main causes of this disease is the lack of vitamin D in the body[40], as well as the type of clothing worn by women in Islamic countries such as Iran, from which the data collection was extracted, has more emphasis on this issue.

Finally, it can be said that the data presented in this article meet most of the ideal criteria. This data has a suitable volume, some personal information of the patient and pictures taken along with their annotations. This information seems to be sufficient for use and generalization. In other words, it can be said that every saved image has metadata and an ID.

## 5. Conclusions

In this study, we introduce an expansive dataset meticulously crafted to address the intricate complexities surrounding osteoporosis, presenting profound implications for the advancement of research, knowledge enrichment, and analytical innovation. Stemming from the meticulous aggregation of DXA bone density measurements, this dataset embodies a multifaceted essence, intertwining crucial individual data intricately with corresponding test outcomes, complemented by meticulously preserved imagery pivotal to these findings, thus enabling thorough analysis and exploration. Beyond its immediate utility, our initiative harbors a broader vision aimed at proactive engagement in combatting the escalating threat posed by osteoporosis. At its core, the provision of this dataset seeks to preemptively address the impending challenges posed by osteoporosis, thereby serving as a potent catalyst for unlocking profound insights into its multifaceted dimensions. Positioned at the forefront of combating this debilitating condition, our dataset holds the promise of yielding monumental benefits for forthcoming generations, arming them with the necessary tools to confront its challenges head-on. Furthermore, the transformative potential inherent in integrating patients’ geographical data into this dataset signifies a critical leap forward. This integration not only empowers spatial analysis but also facilitates the formulation of targeted prevention strategies grounded in spatial data insights, poised to revolutionize the landscape of osteoporosis prevention and treatment. These strategies, fortified by spatial analysis, herald the dawn of a new era in precision medicine, tailored to the unique needs of individuals.

## Data Availability

All data produced are available online athttps://drive.google.com/drive/folders/1HmLTG4GFgB2s4D0x7TTRx8vV_VWY3sW3?usp=sharing

## Acknowledgment

This work was supported in part by the ITRC Support Program under Grant IITP-2024-RS-2022-00156354, in part by the Metaverse Support Program to Nurture the Best Talents under Grant IITP-2024-RS-2023-00254529 funded by the Ministry of Science and ICT of Korea and the Institute of Information and Communications Technology Planning and Evaluation (IITP).

## Notes

### Competing Interest Statement

The authors have declared no competing interest.

### Funding Statement

-

### Author Declarations

Central to this ethical framework was the rigorous scrutiny and approval process facilitated by the ethics committee of the Arak Bone Density Measurement Center

